# Performance of Open-Source LLMs in Challenging Radiological Cases – A Benchmark Study on 1,933 Eurorad Case Reports

**DOI:** 10.1101/2024.09.04.24313026

**Authors:** Su Hwan Kim, Severin Schramm, Lisa C. Adams, Rickmer Braren, Keno K. Bressem, Matthias Keicher, Claus Zimmer, Dennis M. Hedderich, Benedikt Wiestler

## Abstract

Recent advancements in large language models (LLMs) have created new ways to support radiological diagnostics. While both open-source and proprietary LLMs can address privacy concerns through local or cloud deployment, open-source models provide advantages in continuity of access, and potentially lower costs. In this study, we evaluated the diagnostic performance of eleven state-of-the-art open-source LLMs using clinical and imaging descriptions from 1,933 case reports in the Eurorad library. LLMs provided differential diagnoses based on clinical history and imaging findings. Responses were considered correct if the true diagnosis was included in the top three LLM suggestions. Llama-3-70B evaluated LLM responses, with its accuracy validated against radiologist ratings in a case subset. Models were further tested on 60 non-public brain MRI cases from a tertiary hospital to assess generalizability. Llama-3-70B demonstrated superior performance, followed by Gemma-2-27B and Mixtral-8x-7B. Similar performance results were found in the non-public dataset, where Llama-3-70B, Gemma-2-27B, and Mixtral-8x-7B again emerged as the top models. Our findings highlight the potential of open-source LLMs as decision support tools for radiological differential diagnosis in challenging, real-world cases.

## Introduction

Recent advancements in artificial intelligence (AI) have transformed medical diagnostics, offering innovative tools to support clinical decision-making. One promising development is the emergence of large language models (LLMs), which excel at processing and generating natural language. In radiology, these models have demonstrated potential in various applications, including defining study protocols ^1,2^, performing differential diagnosis ^3,4^, generating reports ^5,6^, and extracting information from free-text reports ^7,8^.

However, a significant barrier to the widespread clinical adoption is data privacy. The LLMs primarily used in previous studies are proprietary, closed-source models, such as GPT-4, Claude 3, or Gemini ^9–11^. Access to these models is typically provided via web-based interfaces or via application programming interfaces (API), both of which necessitate the transfer of data to third-party servers, thereby increasing the risk of unauthorized access or misuse of sensitive health information and limiting their use on patient data. While cloud-based solutions for proprietary LLMs can address some privacy concerns, they may still be subject to commercial update cycles and potentially higher long-term costs.

Open-source models offer a viable alternative enabling care institutions to retain patient data within their local infrastructure, mitigating these privacy concerns and providing continuity of access independent of commercial update cycles, which can lower costs due to their free availability. While historically open-source LLMs have underperformed in clinical decision support tasks ^12,13^, Meta’s latest Llama-3 has shown performance levels on par with leading proprietary models in some areas, such as answering radiology board exam questions ^14^. However, the diagnostic accuracy of such models in real-world clinical cases remains largely unexplored.

A well-suited resource for such an evaluation is Eurorad, a comprehensive repository of peer-reviewed radiological case reports managed by the European Society of Radiology (ESR). Eurorad serves as a valuable educational resource for radiologists, residents, and medical students, and encompasses a wide range of cases across radiological subspecialties such as abdominal imaging, neuroradiology, uroradiology and pediatric radiology ^15^.

Therefore, the aim of this study was to evaluate the performance of state-of-the-art open-source LLMs in radiological diagnosis using Eurorad case reports.

## Methods

### Data

To create a comprehensive and diverse dataset of challenging radiology cases, we automatically downloaded case report data—including “Clinical History,” “Imaging Findings,” “Final Diagnosis,” and “Section”—from the European Society of Radiology’s case report library at https://eurorad.org/. All case reports published after July 6, 2015 and licensed under the Creative Commons License CC BY-NC-SA 4.0, were scraped using the Python library “Scrapy” (version 2.11.2) on June 15, 2024.

To address potential data contamination concerns and assess generalizability, we further validated the performance of LLMs in a local dataset of 60 brain MRI cases. These were obtained from our local imaging database, as reported previously ^4^, and equally contained a brief clinical history and imaging findings. This local dataset is not publicly accessible and thus highly unlikely to have been included in the LLMs’ training data.

### LLM Setup

To evaluate a range of open-source large language models (LLMs), we developed a Python-based workflow utilizing the “llama_cpp_python” library (version 0.2.79). This library provides Python bindings for the widely-used “llama_cpp” software, enabling the execution of local, quantized LLMs in GGUF (GPT-Generated Unified Format). Quantization involves reducing the precision of the model’s numerical weights, typically transitioning from floating-point to lower-bit representations, which results in a smaller and faster model while preserving performance. For most models, Q5_K_M was chosen as a quantization, typically offering a good balance between compression and quality. For the 70B models, a quantization factor of Q4_K_M was selected to allow full GPU offloading.

The “llama_cpp_python” library allows for detailed control over relevant hyperparameters. In our experiments, we fully offloaded the LLMs to a GPU for higher computational speed, set the temperature to 0 to ensure deterministic responses, and limited the context width to 1024 tokens, which we previously validated to accommodate all case reports and responses. We chose these settings to balance performance and reproducibility, although we acknowledge that different configurations might yield varying results. Our Python code for prompt construction, along with detailed links to all models (downloaded from https://huggingface.co/), is publicly available in our GitHub repository at https://github.com/ai-idt/os_llm_eurorad.

For this study, we included eleven open-source LLM models, which are detailed in Table 1. All experiments were conducted using an Nvidia P8000 GPU with 48GB of video memory.

**Table 1:** Model details.

| Model | No. of Parameters | Link to Base Model |
| --- | --- | --- |
| Meta-Llama-3-70B | 70 billion | https://huggingface.co/meta-llama/Meta-Llama-3-70B-Instruct |
| Gemma-2-27B | 27 billion | https://huggingface.co/google/gemma-2-27b-it |
| Mixtral-8x7B | 47 billion | https://huggingface.co/mistralai/Mixtral-8x7B-Instruct-v0.1 |
| Meta-Llama-3-8B | 8 billion | https://huggingface.co/meta-llama/Meta-Llama-3-8B-Instruct |
| Phi-3-medium-128K | 14 billion | https://huggingface.co/microsoft/Phi-3-medium-128k-instruct |
| Llama-2-70B | 70 billion | https://huggingface.co/meta-llama/Llama-2-70b-chat-hf |
| Vicuna-13B | 13 billion | https://huggingface.co/lmsys/vicuna-13b-v1.5 |
| BioMistral-7B | 7 billion | https://huggingface.co/BioMistral/BioMistral-7B |
| Meditron-7B | 7 billion | https://huggingface.co/epfl-llm/meditron-7b |
| OpenBioLLM-Llama3-8B | 8 billion | https://huggingface.co/aaditya/Llama3-OpenBioLLM-8B |
| Medalpaca-13B | 13 billion | https://huggingface.co/medalpaca/medalpaca-13b |

### Case Selection and Response Assessment

Upon review, we noted that a significant proportion of cases already contained the correct diagnosis within the “Clinical History” and “Imaging Findings” sections. Drawing inspiration from the “LLM-as-a-Judge” paradigm ^16^, we employed the most advanced model available at the outset of this study, Llama-3-70B, to filter out these cases. A recent study indicated that Llama-3-70B, along with GPT-4 Turbo, demonstrated the closest alignment with human evaluations ^17^, making it particularly suitable for this task. We prompted Llama-3-70B to assess all cases with the following instruction:

“*You are a senior radiologist. Below, you will find a case description for a patient diagnosed with [Diagnosis]. Please check if the diagnosis or any part of it is mentioned, discussed, or suggested in the case description. Respond with either ‘mentioned’ (if the diagnosis is included) or ‘not mentioned*,*’ and nothing else*.”

Subsequently, we prompted each of the eleven LLMs to provide three differential diagnoses along with a brief rationale for each, using the concatenated “Clinical History” and “Imaging Findings” as input:

“*You are a senior radiologist. Below, you will find information about a patient: first, the clinical presentation, followed by imaging findings. Based on this information, name the three most likely differential diagnoses, with a short rationale for each*.”

Finally, we again utilized Llama-3-70B to evaluate each LLM’s responses on a binary scale, categorizing them as either “correct” (if the correct diagnosis was among the three differential diagnoses) or “wrong.” The prompt for this evaluation was:

“*You are a senior radiologist. Below, you will find the correct diagnosis (indicated after ‘Correct Diagnosis:’) followed by the differential diagnoses provided by a Radiology Assistant during an exam. Please assess whether the Radiology Assistant included the correct diagnosis in their differential diagnosis. Respond only with ‘correct’ (if the correct diagnosis is included) or ‘wrong’ (if it is not)*.”

### Human Evaluation

In order to gain an understanding of Llama-3-70B’s performance as an LLM judge for correctness of diagnoses, three experienced radiologists (SHK, with 2 years of experience, DMH and BW, board-certified radiologists with 10 years of experience each) additionally evaluated 60 LLM responses each for correctness, of which 20 were shared between all three reviewers to assess human inter-rater agreement. Using a total of 140 LLM responses for which both human “ground truth” and LLM judge assessments were known, we calculated the accuracy of the LLM judge (Figure 1).

**Figure 1.**
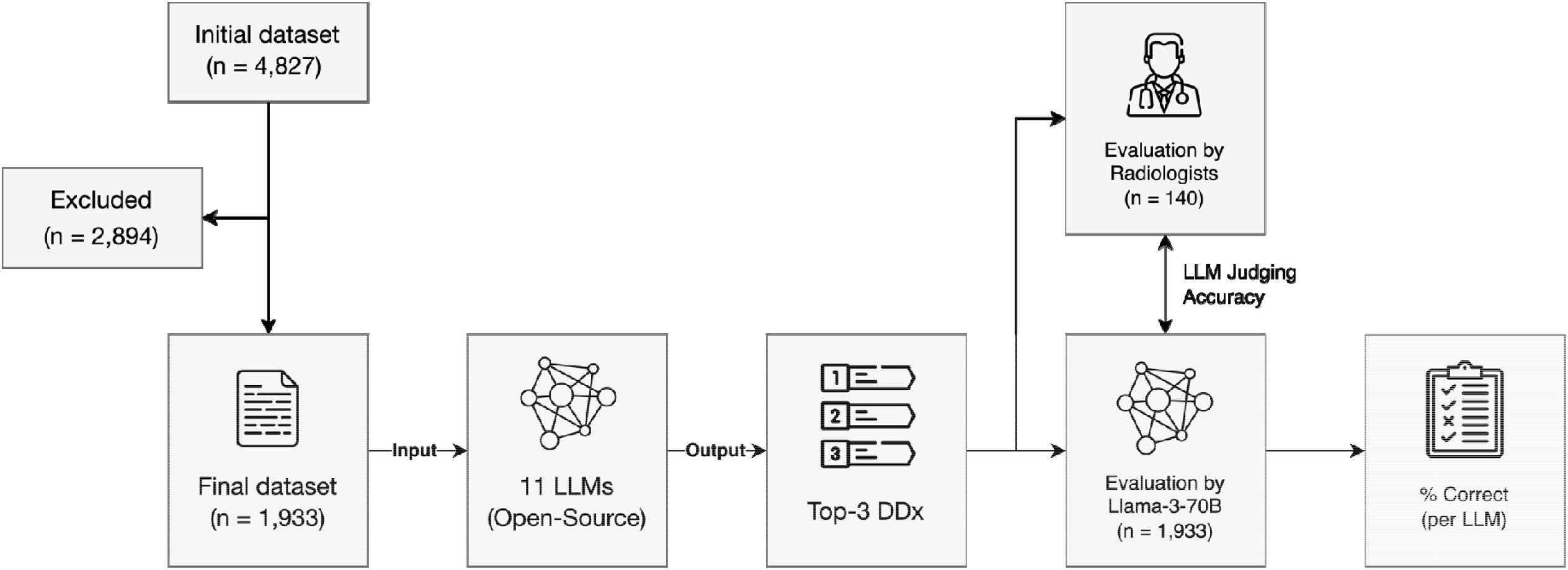
Study Design. A total of 2,894 cases were excluded as the true diagnosis was mentioned in the case description to be provided as LLM input. DDx: differential diagnoses.

### Statistics

Both the LLM judge as well as human raters evaluated LLM responses on a binary scale, i.e., if the correct diagnosis was among the top 3 differential diagnoses listed by the LLM or not. From this response data, we calculated the standard error per model and category as:

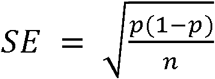

where *p* is the proportion of correct responses, and *n* is the number of samples. However, from our human evaluation of the LLM judge performance, we know about its inaccuracies and have to adjust the SE to account for this:

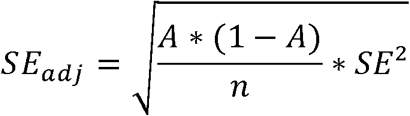

where *A* is the accuracy of the LLM judge. The adjusted 95% Confidence Interval is then:

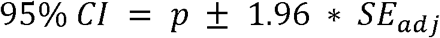

## Results

### Dataset

The initial dataset retrieved from the Eurorad library consisted of 4,827 case reports. Using the Llama-3-70B model, we identified 2,894 cases where the diagnosis was explicitly stated within the case description. These cases were subsequently excluded, resulting in a final dataset of 1,933 cases for analysis. This filtering process ensured that the LLMs were evaluated on genuinely challenging cases that required inference rather than simple information extraction. The dataset was primarily composed of cases from neuroradiology (21.4%), abdominal imaging (18.1%), and musculoskeletal imaging (14.6%), whereas breast imaging (3.4%) and interventional radiology (1.4%) were underrepresented (Table 2). This distribution broadly reflects the relative prevalence of different radiological subspecialties in clinical practice.

**Table 2:** Dataset composition by subspecialty.

| Subspecialty | No. Cases | Proportion |
| --- | --- | --- |
| Neuroradiology | 413 | 21.4% |
| Abdominal imaging | 349 | 18.1% |
| Musculoskeletal system | 282 | 14.6% |
| Chest imaging | 190 | 9.8% |
| Uroradiology & genital male imaging | 151 | 7.8% |
| Paediatric radiology | 137 | 7.1% |
| Head & neck imaging | 124 | 6.4% |
| Cardiovascular | 112 | 5.8% |
| Genital (female) imaging | 83 | 4.3% |
| Breast imaging | 65 | 3.4% |
| Interventional radiology | 27 | 1.4% |
| Total | 1933 | 100.0% |

### LLM Judge Performance

Based on 140 LLM responses rated by radiologists as the reference standard, Llama-3-70B exhibited an accuracy of 87.8% in classifying responses as “correct” or “incorrect” (123/140 responses; 95% CI: 0.82 – 0.93). Furthermore, in a subset of 20 responses rated by all three radiologists, the interrater agreement was found to be 100%, indicating complete consensus. This high level of agreement between Llama-3-70B and human radiologists, as well as among radiologists themselves, supports the validity of using Llama-3-70B as an automated judge for the larger dataset.

### Model Performance

Across all models, the highest levels of diagnostic accuracy were achieved in interventional radiology (65.7 ± 7.3%), cardiovascular imaging (58.8 ± 3.7%), and abdominal imaging (57.0 ± 2.1%), whereas lower accuracy was observed in breast imaging (48.2 ± 5.0%) and musculoskeletal imaging (47.3 ± 2.4%) (Figure 2). Granular accuracy metrics by subspecialty and model are provided in Supplement 1. Among the evaluated models, Llama-3-70B demonstrated superior diagnostic performance across all subspecialties, achieving a rate of 73.2 ± 2.5% correct responses, a considerable margin ahead of Gemma-2-27B (62.4 ± 2.6%), Mixtral-8x7B (57.8 ± 2.6%), and Meta-Llama-3-8B (56.4 ± 2.6%) (Figure 3).

**Figure 2.**
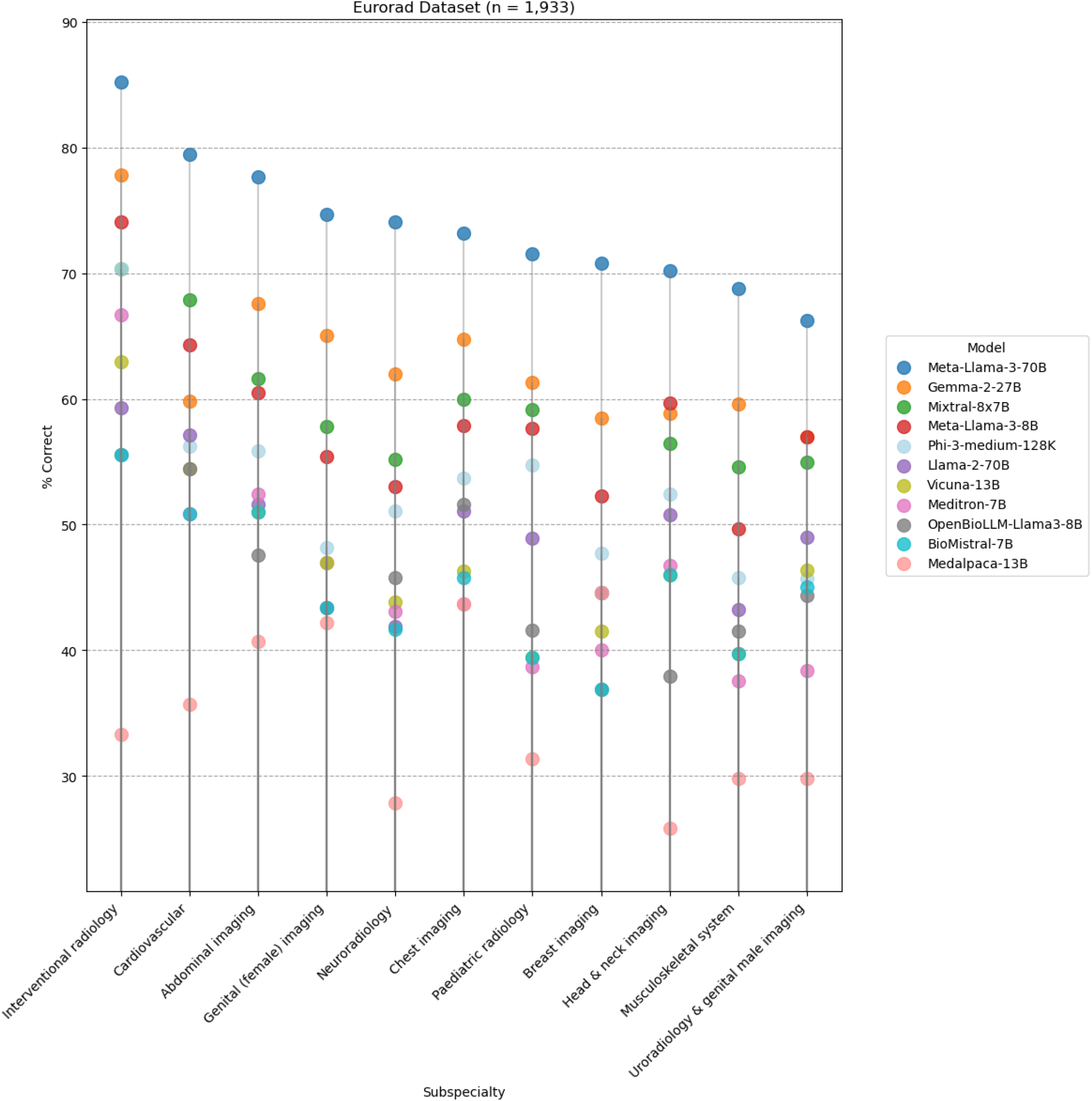
Model Performance by Subspecialty. Meta-Llama-3-70B demonstrated highest performance across all subspecialties.

**Figure 3.**
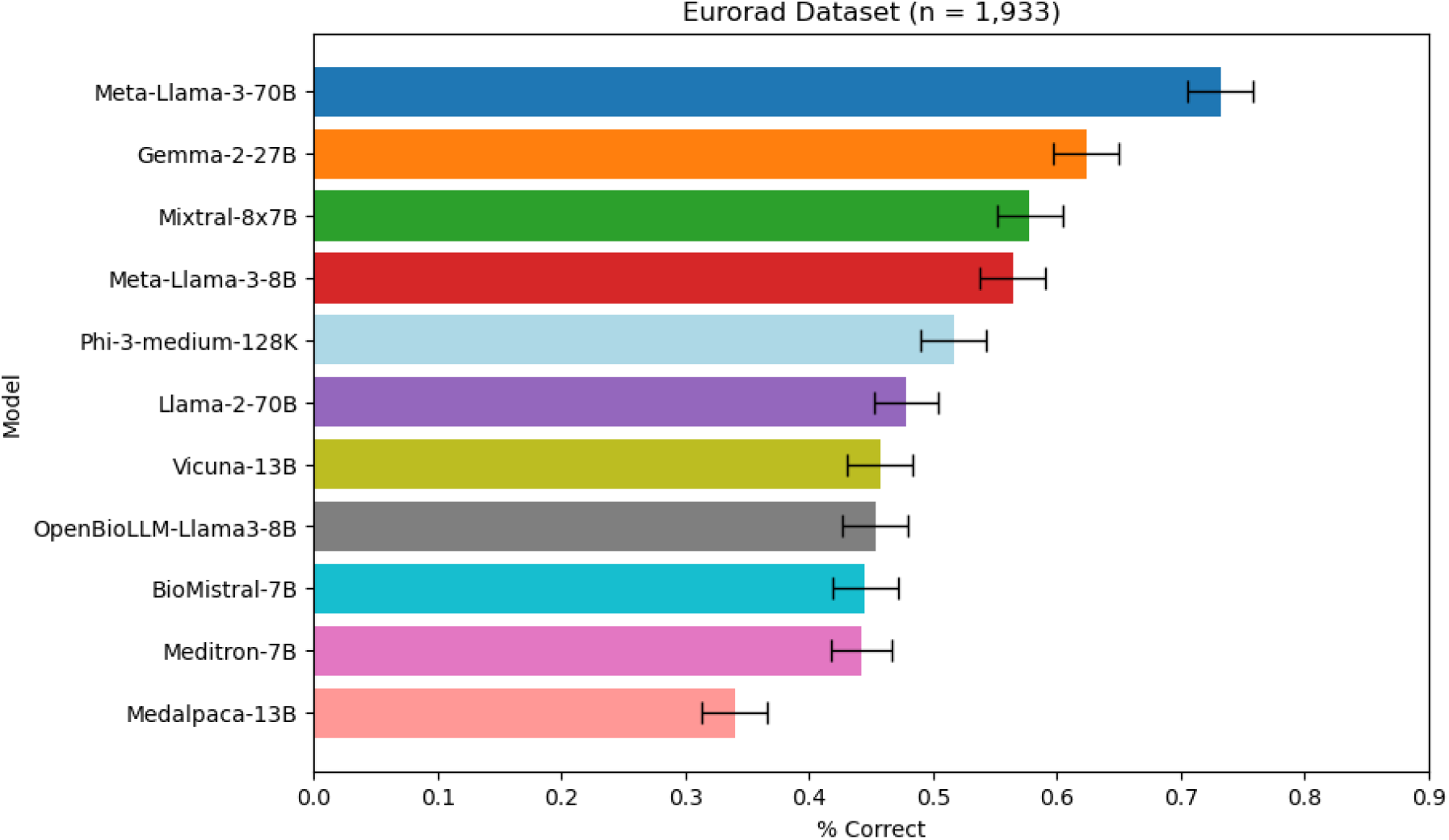
Performance of Open-Source LLMs in Eurorad dataset (n = 1,933). Error bars indicate adjusted 95% confidence intervals.

In the local brain MRI dataset, comparable results were observed, with Llama-3-70B (71.7 ± 14.1%), Gemma-2-27B (53.3 ± 15.1%), and Mixtral-8x-7B (51.7 ± 15.1%) again leading the rankings (Figure 4). The consistent performance on this non-public dataset suggests that the models’ capabilities generalize beyond potentially contaminated public data, reinforcing the robustness of our findings.

**Figure 4.**
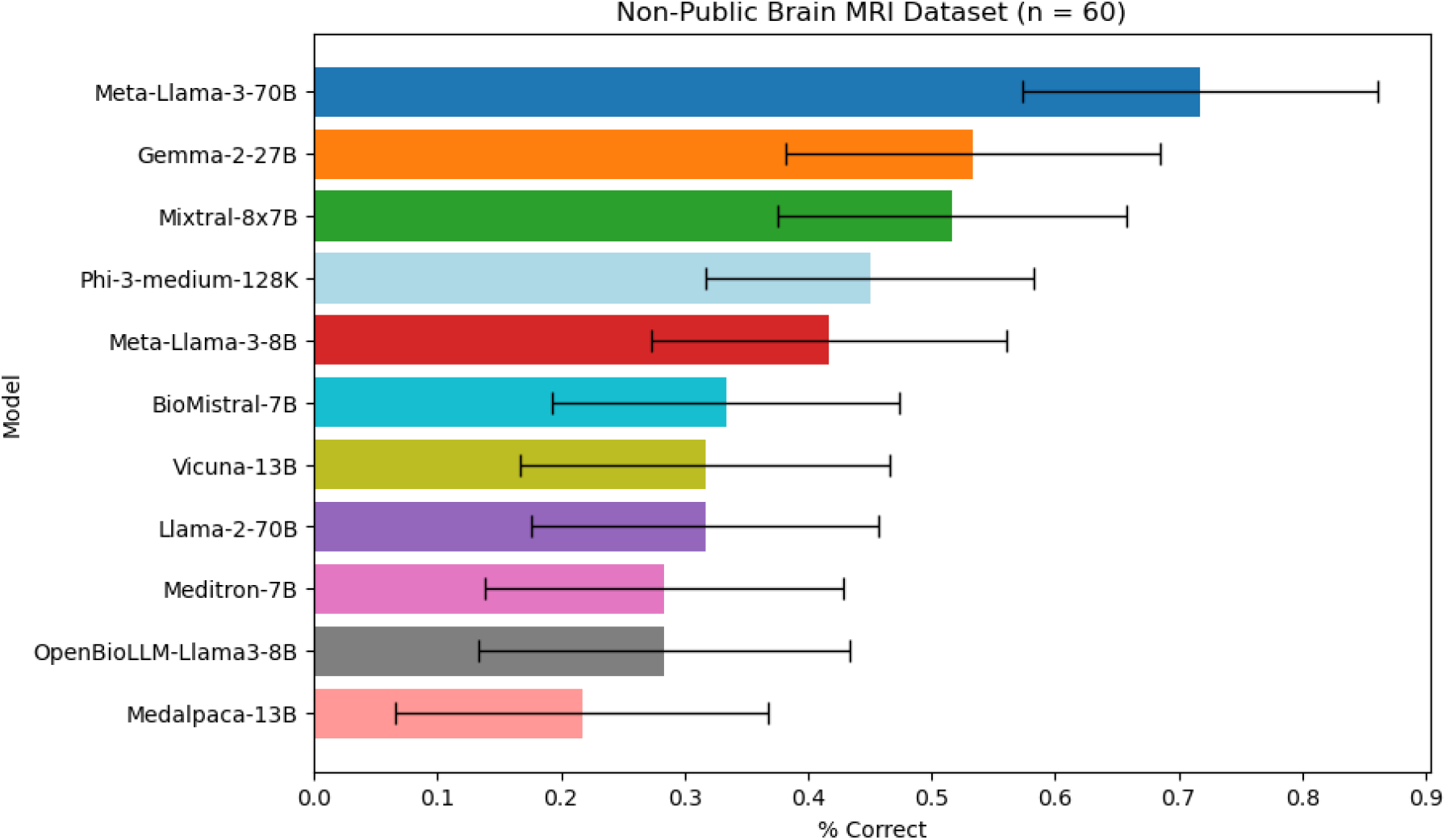
Performance of Open-Source LLMs in non-public brain MRI dataset (n = 60). Error bars indicate adjusted 95% confidence intervals.

## Discussion

In this study, we benchmarked the diagnostic performance of eleven leading open-source LLMs in a heterogeneous, challenging cohort of 1,933 peer-reviewed case reports from the Eurorad library. Meta’s Llama-3-70B demonstrated superior performance, surpassing the other models across all radiological subspecialties with an overall accuracy of 73.2%. This level of performance is particularly noteworthy given the complexity and diversity of the cases included in our dataset.

These findings underscore the current dominance of Llama-3 among open-source models, consistent with its proficiency in other clinical tasks, such as answering close-ended medical questions, summarizing clinical documents, and patient education ^14,18^.

Importantly, this study assessed the diagnostic performance of LLMs based on real case descriptions, more accurately representing the complexities of real-life clinical decision-making than questions with pre-defined response options. This approach provides a more realistic evaluation of LLMs’ potential in clinical settings, where the ability to interpret nuanced clinical information is crucial.

Our results revealed interesting variations in performance across radiological subspecialties, with higher accuracy in genital (female) imaging and lower accuracy in musculoskeletal imaging. These differences may reflect inherent complexities within each subspecialty, variations in the quality or specificity of case descriptions, or potential biases in the models’ training data. Further investigation into these subspecialty-specific performance variations could provide valuable insights for targeted model improvements and clinical applications.

Interestingly, some lighter models such as Meta-Llama-3-8B exhibited strong performance, outperforming larger models with more parameters (e.g. Llama-2-70B, Vicuna-13B). This suggests that smaller, lower-cost models with nonetheless robust results are attainable, making the implementation of LLMs in resource-constrained healthcare settings more viable. The strong performance of smaller models highlights the importance of model architecture and training strategies, rather than just model size, in achieving high performance on specialized tasks.

Employing a state-of-the-art LLM model to automate the evaluation of LLM responses facilitated the large-scale analysis of thousands of cases, a scope unrealizable through manual processing. This strategy establishes a methodical benchmark for future large-scale investigations of clinical text documents.

### Limitations

First, data contamination of LLMs cannot be definitively ruled out. Given the lack of transparency regarding the LLM training datasets, it is possible that the case reports used in this study overlap with the training data of some models. However, our complementary assessment on a non-public brain MRI dataset revealed only a minor drop in performance, while the overall model rankings remained nearly identical.

Second, while the use of an LLM for the evaluation of LLM responses significantly enhanced the scalability of the analysis, it did so at the expense of reduced accuracy. To mitigate this limitation, we adjusted the standard error of model performance assessment based on our evaluation of Llama-3-70B’s judging accuracy in a subset of the data.

Third, we did not investigate the impact of temperature settings or prompt design on LLM performance. To ensure deterministic responses, we applied a temperature of 0, but higher temperatures could potentially improve diagnostic accuracy ^10^. Similarly, the optimal task-specific prompting strategy for radiological diagnosis is yet to be determined ^19^.

Finally, this study did not account for the influence of varying descriptions of the same case. A recent study evaluating GPT-4(V) in radiological diagnosis revealed that the image description is a major determinant of LLM accuracy ^4^. The Eurorad case descriptions were written in awareness of the correct diagnosis, and their use of specific terminology or emphasis on certain image characteristics might have introduced a positive bias in LLM performance.

In conclusion, we found that several open-source LLMs demonstrate promising performance in identifying the correct diagnosis based on case descriptions from the Eurorad library, highlighting their potential as decision support tool for radiological differential diagnosis.

## Data Availability

The Python code used in the present study is publicly available online at our GitHub repository (https://github.com/ai-idt/os_llm_eurorad).

https://eurorad.org/

https://github.com/ai-idt/os_llm_eurorad

