## Supplement 1 for "Performance of Open-Source LLMs in Challenging Radiological Cases – A Benchmark Study on 1,933 Eurorad Case Reports"

| Subspecialty | Model | % Correct | CI_lower | CI_upper |
| --- | --- | --- | --- | --- |
| Abdominal imaging | Meta-Llama-3-70B | 77.7% | 72.1% | 83.2% |
|  | Gemma-2-27B | 67.6% | 61.6% | 73.6% |
|  | Mixtral-8x7B | 61.6% | 55.5% | 67.8% |
|  | Meta-Llama-3-8B | 60.5% | 54.3% | 66.6% |
|  | Phi-3-medium-128K | 55.9% | 49.6% | 62.1% |
|  | Meditron-7B | 52.4% | 46.2% | 58.7% |
|  | Llama-2-70B | 51.6% | 45.3% | 57.8% |
|  | BioMistral-7B | 51.0% | 44.7% | 57.3% |
|  | Vicuna-13B | 51.0% | 44.7% | 57.3% |
|  | OpenBioLLM-Llama3-8B | 47.6% | 41.3% | 53.8% |
|  | Medalpaca-13B | 40.7% | 34.5% | 46.9% |
| Breast imaging | Meta-Llama-3-70B | 70.8% | 57.2% | 84.4% |
|  | Gemma-2-27B | 58.5% | 44.1% | 72.8% |
|  | Meta-Llama-3-8B | 52.3% | 37.8% | 66.8% |
|  | Phi-3-medium-128K | 47.7% | 33.2% | 62.2% |
|  | Llama-2-70B | 44.6% | 30.2% | 59.1% |
|  | Medalpaca-13B | 44.6% | 30.2% | 59.1% |
|  | Mixtral-8x7B | 44.6% | 30.2% | 59.1% |
|  | Vicuna-13B | 41.5% | 27.2% | 55.9% |
|  | Meditron-7B | 40.0% | 25.7% | 54.3% |
|  | BioMistral-7B | 36.9% | 22.8% | 51.1% |
|  | OpenBioLLM-Llama3-8B | 36.9% | 22.8% | 51.1% |
| Cardiovascular | Meta-Llama-3-70B | 79.5% | 69.8% | 89.1% |
|  | Mixtral-8x7B | 67.9% | 57.3% | 78.4% |
|  | Meta-Llama-3-8B | 64.3% | 53.5% | 75.0% |
|  | Gemma-2-27B | 59.8% | 48.9% | 70.7% |
|  | Llama-2-70B | 57.1% | 46.2% | 68.1% |
|  | Phi-3-medium-128K | 56.3% | 45.2% | 67.3% |
|  | OpenBioLLM-Llama3-8B | 54.5% | 43.4% | 65.5% |
|  | Vicuna-13B | 54.5% | 43.4% | 65.5% |
|  | BioMistral-7B | 50.9% | 39.8% | 62.0% |
|  | Meditron-7B | 50.9% | 39.8% | 62.0% |
|  | Medalpaca-13B | 35.7% | 25.0% | 46.5% |
| Chest imaging | Meta-Llama-3-70B | 73.2% | 65.3% | 81.0% |
|  | Gemma-2-27B | 64.7% | 56.5% | 73.0% |
|  | Mixtral-8x7B | 60.0% | 51.6% | 68.4% |
|  | Meta-Llama-3-8B | 57.9% | 49.5% | 66.3% |
|  | Phi-3-medium-128K | 53.7% | 45.2% | 62.2% |
|  | OpenBioLLM-Llama3-8B | 51.6% | 43.1% | 60.1% |
|  | Llama-2-70B | 51.1% | 42.6% | 59.5% |
|  | Vicuna-13B | 46.3% | 37.8% | 54.8% |
|  | BioMistral-7B | 45.8% | 37.3% | 54.3% |
|  | Medalpaca-13B | 43.7% | 35.2% | 52.1% |
|  | Meditron-7B | 43.7% | 35.2% | 52.1% |
| Genital (female) imaging | Meta-Llama-3-70B | 74.7% | 63.0% | 86.4% |
|  | Gemma-2-27B | 65.1% | 52.6% | 77.5% |
|  | Mixtral-8x7B | 57.8% | 45.1% | 70.6% |
|  | Meta-Llama-3-8B | 55.4% | 42.6% | 68.2% |
|  | Phi-3-medium-128K | 48.2% | 35.3% | 61.0% |
|  | Llama-2-70B | 47.0% | 34.2% | 59.8% |
|  | Vicuna-13B | 47.0% | 34.2% | 59.8% |
|  | BioMistral-7B | 43.4% | 30.6% | 56.1% |
|  | Meditron-7B | 43.4% | 30.6% | 56.1% |
|  | OpenBioLLM-Llama3-8B | 43.4% | 30.6% | 56.1% |
|  | Medalpaca-13B | 42.2% | 29.4% | 54.9% |
| Head & neck imaging | Meta-Llama-3-70B | 70.2% | 60.3% | 80.1% |
|  | Meta-Llama-3-8B | 59.7% | 49.3% | 70.1% |
|  | Gemma-2-27B | 58.9% | 48.5% | 69.3% |
|  | Mixtral-8x7B | 56.5% | 46.0% | 66.9% |
|  | Phi-3-medium-128K | 52.4% | 41.9% | 62.9% |
|  | Llama-2-70B | 50.8% | 40.3% | 61.3% |
|  | Meditron-7B | 46.8% | 36.3% | 57.3% |
|  | BioMistral-7B | 46.0% | 35.5% | 56.5% |
|  | Vicuna-13B | 46.0% | 35.5% | 56.5% |
|  | OpenBioLLM-Llama3-8B | 37.9% | 27.6% | 48.2% |
|  | Medalpaca-13B | 25.8% | 16.2% | 35.4% |
| Interventional radiology | Meta-Llama-3-70B | 85.2% | 67.0% | 103.4% |
|  | Gemma-2-27B | 77.8% | 57.8% | 97.7% |
|  | Meta-Llama-3-8B | 74.1% | 53.5% | 94.7% |
|  | Mixtral-8x7B | 70.4% | 49.2% | 91.5% |
|  | Phi-3-medium-128K | 70.4% | 49.2% | 91.5% |
|  | Meditron-7B | 66.7% | 45.0% | 88.3% |
|  | Vicuna-13B | 63.0% | 41.0% | 85.0% |
|  | Llama-2-70B | 59.3% | 37.0% | 81.5% |
|  | BioMistral-7B | 55.6% | 33.1% | 78.0% |
|  | OpenBioLLM-Llama3-8B | 55.6% | 33.1% | 78.0% |
|  | Medalpaca-13B | 33.3% | 11.7% | 55.0% |
| Musculoskeletal system | Meta-Llama-3-70B | 68.8% | 62.2% | 75.4% |
|  | Gemma-2-27B | 59.6% | 52.7% | 66.5% |
|  | Mixtral-8x7B | 54.6% | 47.7% | 61.6% |
|  | Meta-Llama-3-8B | 49.6% | 42.7% | 56.6% |
|  | Phi-3-medium-128K | 45.7% | 38.8% | 52.7% |
|  | Llama-2-70B | 43.3% | 36.3% | 50.2% |
|  | OpenBioLLM-Llama3-8B | 41.5% | 34.6% | 48.4% |
|  | BioMistral-7B | 39.7% | 32.8% | 46.6% |
|  | Vicuna-13B | 39.7% | 32.8% | 46.6% |
|  | Meditron-7B | 37.6% | 30.8% | 44.4% |
|  | Medalpaca-13B | 29.8% | 23.2% | 36.3% |
| Neuroradiology | Meta-Llama-3-70B | 74.1% | 68.8% | 79.4% |
|  | Gemma-2-27B | 62.0% | 56.3% | 67.6% |
|  | Mixtral-8x7B | 55.2% | 49.5% | 60.9% |
|  | Meta-Llama-3-8B | 53.0% | 47.3% | 58.8% |
|  | Phi-3-medium-128K | 51.1% | 45.3% | 56.8% |
|  | OpenBioLLM-Llama3-8B | 45.8% | 40.0% | 51.5% |
|  | Vicuna-13B | 43.8% | 38.1% | 49.6% |
|  | Meditron-7B | 43.1% | 37.4% | 48.8% |
|  | Llama-2-70B | 41.9% | 36.2% | 47.6% |
|  | BioMistral-7B | 41.6% | 35.9% | 47.3% |
|  | Medalpaca-13B | 27.8% | 22.5% | 33.2% |
| Paediatric radiology | Meta-Llama-3-70B | 71.5% | 62.2% | 80.9% |
|  | Gemma-2-27B | 61.3% | 51.5% | 71.1% |
|  | Mixtral-8x7B | 59.1% | 49.2% | 69.0% |
|  | Meta-Llama-3-8B | 57.7% | 47.7% | 67.6% |
|  | Phi-3-medium-128K | 54.7% | 44.8% | 64.7% |
|  | Llama-2-70B | 48.9% | 38.9% | 58.9% |
|  | OpenBioLLM-Llama3-8B | 41.6% | 31.7% | 51.5% |
|  | BioMistral-7B | 39.4% | 29.6% | 49.3% |
|  | Vicuna-13B | 39.4% | 29.6% | 49.3% |
|  | Meditron-7B | 38.7% | 28.9% | 48.5% |
|  | Medalpaca-13B | 31.4% | 21.9% | 40.9% |
| Uroradiology & genital male imaging | Meta-Llama-3-70B | 66.2% | 57.1% | 75.4% |
|  | Gemma-2-27B | 57.0% | 47.5% | 66.4% |
|  | Meta-Llama-3-8B | 57.0% | 47.5% | 66.4% |
|  | Mixtral-8x7B | 55.0% | 45.5% | 64.5% |
|  | Llama-2-70B | 49.0% | 39.5% | 58.5% |
|  | Vicuna-13B | 46.4% | 36.8% | 55.9% |
|  | Phi-3-medium-128K | 45.7% | 36.2% | 55.2% |
|  | BioMistral-7B | 45.0% | 35.5% | 54.5% |
|  | OpenBioLLM-Llama3-8B | 44.4% | 34.9% | 53.9% |
|  | Meditron-7B | 38.4% | 29.1% | 47.8% |
|  | Medalpaca-13B | 29.8% | 20.8% | 38.8% |

Supplement 1: Accuracy with 95% confidence intervals by subspecialty and model (Eurorad dataset).
